# A new tridimensional effective method to calculate Real Contact Surface Area (RCSA) between renal tumor and surrounding kidney parenchyma: An Innovative Tool with predictive power of intraoperative outcomes in nephron sparing surgery

**DOI:** 10.1101/2024.01.12.23295420

**Authors:** Paolo Traverso, Alessandro Carfì, Alessandra Bulanti, Martina Fabbi, Veronica Giasotto, Matilde Mattiauda, Lorenzo Lo Monaco, Stefano Tappero, Giovanni Guano, Federica Balzarini, Marco Borghesi, Fulvio Mastrogiovanni, Carlo Terrone

## Abstract

The Contact Surface Area (CSA) is a predictor for peri-operative parameters and represent the contact area between the tumor and the respective organ. Nowadays, a precise method for calculating CSA is yet to be found in the literature. We tested a new CSA calculation method as a predictor of intra-operative parameters in robot assisted partial nephrectomy (RAPN).

The study population consisted of all consecutive patients treated with RAPN at a single high-volume European institution (between 2020 to 2023; 82 patients). We proposed a new method to measure the real value of CSA using an algorithm that leverages the geometry of kidneys and tumors obtained from 3D reconstruction. These reconstructions were obtained using the certified medical software Materialized Mimics InPrint. Peri-operative parameters of patients were recorded in an anonymous database.

We explored the correlation between RCSA, CSA of Hsieh (HCSA), PADUA and R.E.N.A.L. scores with peri-operative parameters using Spearman’s correlation. Furthermore, we examined which of RCSA, PADUA and R.E.N.A.L. score better describes the intra-operative parameters, Warm Ischemia Time (WIT), Operating Time (OT), and Estimated Blood Loss (EBL) using Receiver Operating Characteristic (ROC) curve analysis. Multivariable linear regression analyses were performed.

We observed a significant correlation between RCSA and WIT, OT and EBL. Moreover, RCSA outperformed both the PADUA and R.E.N.A.L. score as demonstrated in the ROC curve analysis. In ROC analysis was chosen a threshold for each of the parameters: for WIT 20 minutes, for OT 180 minutes and for EBL 200 mL. At multivariable regression analysis, RCSA emerged as the only independent predictor for WIT, OT and EBL (B=0.39 & p=0.03, B=0.35 & p=0.01, B=0.48 & p*<*0.001, respectively).

Our original and effective 3D RCSA calculation method was favorably associated to intra-operative surgical outcomes. As compared to PADUA and RENAL score, our calculated RCSA represented a better predictor of intra-operative parameters.

## 1 Introduction

Renal cell carcinoma (RCC) accounts for 3% of all solid malignancies and ranks tenth in terms of prevalence, with a higher number of cases in western countries [1, 2]. The main objective in the treatment of localized stage I renal masses is to maximally respect the renal parenchyma with surgical precision, avoiding surgical margins [1]. 3D reconstruction plays an important role approaching both easy and complex tumor masses that overturn renal anatomy especially when vascular anatomy is surgically challenging [3, 4].

Several scores have been considered to approach *robot-assisted partial nephrectomy* (RAPN) in order to standardized the characterization of renal tumours and assess the complexity of *Nephron sparing surgery* (NSS), correlating with some perioperative outcomes; pure nephrometry scores (radius [R], exophytic/endophytic [E], nearness to collecting system/sinus [N], anterior/posterior [A], and location relative to polar lines [L] [RENAL], preoperative aspects and dimensions used for an anatomical classification [PADUA], Simplified Padua Renal Nephrectomy [SPARE], C-Index, Diameter-Axial-Polar [DAP], Contact Surface Area [CSA]) [5–9] can provide information on the postoperative course or on intra-operative difficulties. Considering precision surgery, having indexes to provide the most complete overview is essential.

All the scores mentioned above were formulated during a period when 3D reconstruction had not yet been extensively developed and applied. The concept of CSA was introduced by Leslie et al. in 2014 [10] and it is a variable and an anatomical measure that aims to quantify the contact area between the renal tumor and the surrounding healthy parenchyma. Specifically, the CSA calculation was based on the assumption that all kidney tumors were similar to spheres. It reflects two indicative anatomical factors of surgical complexity: the size of the tumor and its degree of intraparenchymal extension [7, 11]. In our study, we reassess the values of CSA in our cohort of patients treated with RAPN, using an innovative calculation system based on 3D DICOM segmentation and we correlate the new contact surfaces area values with perioperative outcomes after RAPN.

## 2 Materials & Methods

### 2.1 Data acquisition

We evaluated all consecutive patients undergoing RAPN between November 2020 to May 2023 at our Institute, “Policlinico San Martino”, operated by the same surgeon. We included 78 patients out of the 82 who underwent RAPN excluding those with more than one tumor or bilateral tumors.

We collected demographic data Body Mass Index (BMI), age and sex. Clinical data included post-operative and follow-up parameters such as haemoglobin (HB), creatinine (CREA) and estimate glomerular filtration rate (eGFR). Intra-operative surgical data comprised warm ischemia time (WIT), operative time (OT) and estimate blood loss (EBL). Pathological features such as histotype (T,N,M), staging and benign/malignant classification were also recorded for each patient. We also considered nephrometry scores as PADUA score and R.E.N.A.L. score.

The measure of eGFR relied on the CKD-EPI equation adjusted for creatinine values [12]. Pre-operative data was recorded one day before the operation, while post-operative data 3 days after the operation. Furthermore follow-up data were recorded 6 months after the operation. Therefore, only patients operated between November 2020 and Dicember 2022 had follow-up data. Additionally, the percentage change in eGFR (PCE) was calculated for each patient using the following formula: *(eGFR post-eGFR pre)/eGFR pre*.

### 2.2 CT Protocol

In case that radiological examinations of the tumor were not adequate and available for the preoperative planning of RAPN, we acquired the images with a SIEMENS SOMATON DEFINITION FLASH 128 scanner with a technique dedicated to 3D reconstructions, which provided a double administration of iodinated contrast agent 10 minutes apart, 50 + 90 ml (split bolus technique) and the execution of an arterial phase scan with bolus tracking (delay 10 sec) acquired after the second administration of contrast agent, at 3.5 mL /sec. This dedicated protocol allows the simultaneous opacification of the urinary tract, the renal arterial and venous vessels by the contrast agent (using the physiological early renal venous opacification).

### 2.3 Segmentation

Not all the images we used for segmentation were acquired with the protocol described above as some patients underwent CT scan in other center. However for all patients we utilized the same phase for segmentation, specifically arterial phase. This phase provided the best visualization of the kidneys and tumors, allowing for accurate recognition.

For the reconstruction of the kidney and tumors, we utilized a certified medical software Materialise Mimics InPrint. The absence of holes in the parenchyma involved in the contact surface is a necessary condition for computing CSA as depicted in Figure 1. We used the reconstructed kidneys and tumors to compute the real value of CSA (Figure 2).

**Figure 1:**
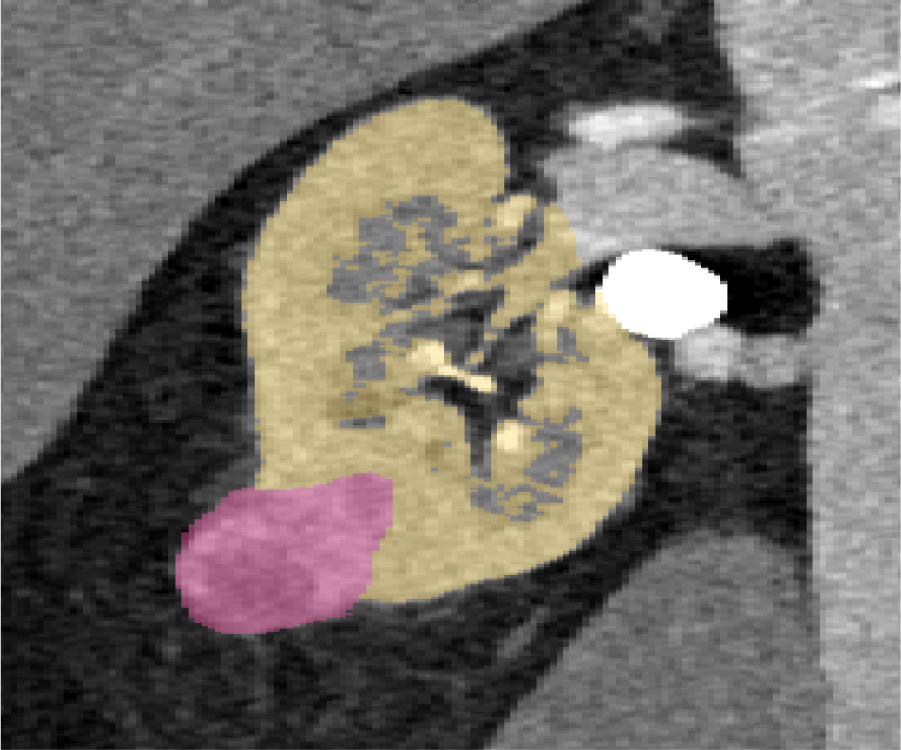
In this image, the tumor is depicted in pink, while the kidney is represented in yellow. As can be seen there are no holes or openings at the interface where the kidney and tumor meet.

**Figure 2:**
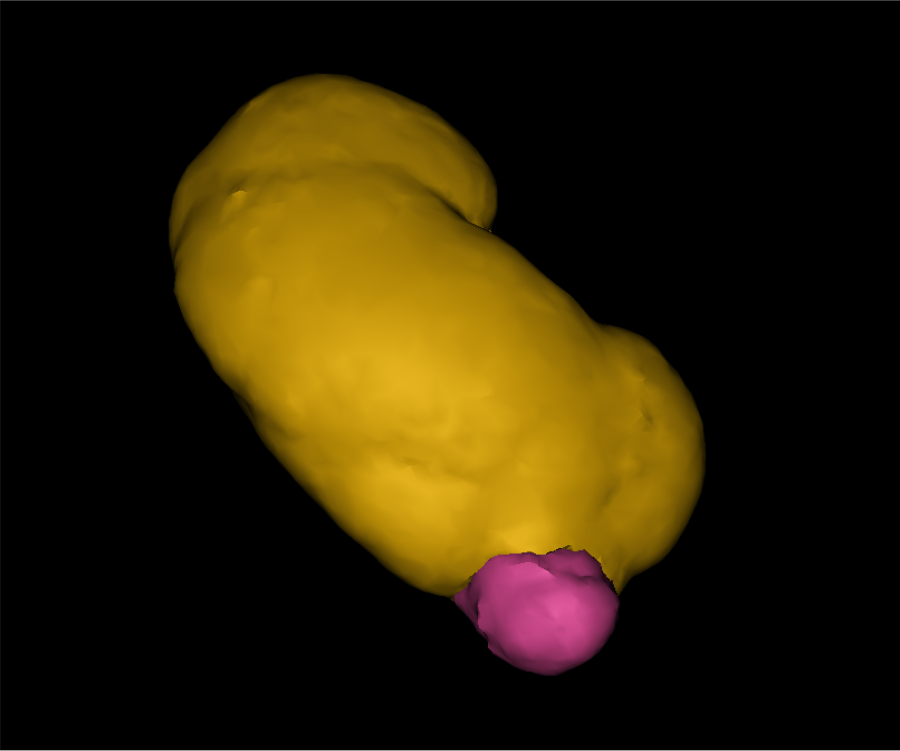
3D reconstruction of kidney and tumor that we extract from Materialise as STL files.

### 2.4 Algorithm

An algorithm was developed that leverages the geometry of the tumor and the organ to estimate the real value of CSA.

Our software processed the 3D reconstructions of the kidney and tumor, uploaded as STL files, and returned the *Real Contact Surface Area* (RCSA) together with a 3D visualization of the tumor and of the RCSA, as shown in Figure 3. Additionally, the algorithm provided the values of the total area and total volume of the tumor. These values were compared with the measurement of the total area and volume automatically computed by Materialise after the 3D reconstruction, and they were found to be consistent.

**Figure 3:**
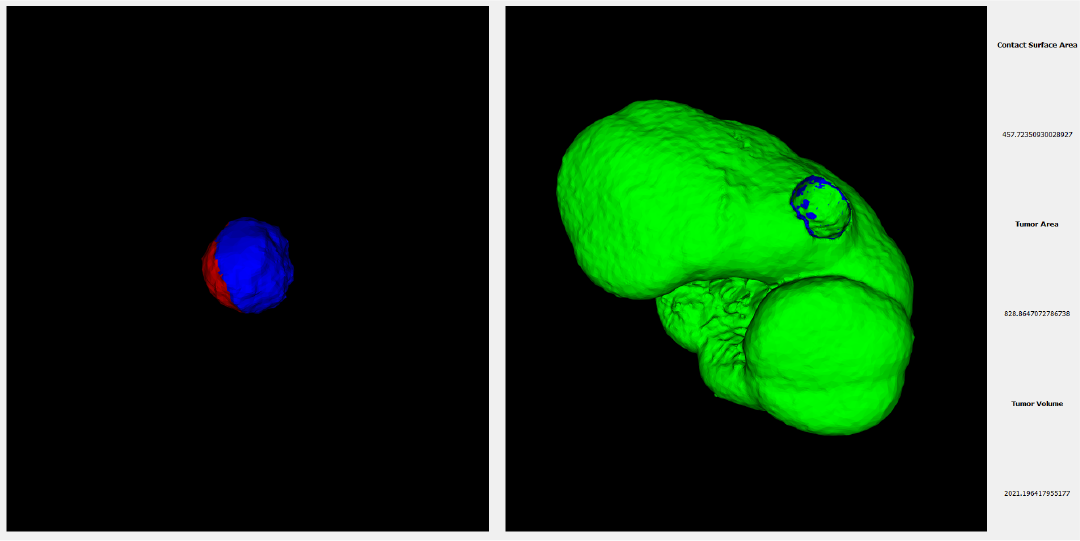
This image illustrates the results of the algorithm: presenting 3D graphs of the tumor and kidney, alongside the measurements of RCSA, Total Tumor Area, and Total Tumor Volume. Notably, when focusing on the tumor, the blue segment signifies the contact area with the kidney.

The performance of the algorithm in estimating the RCSA was evaluated using a synthetic benchmark built using a *Computer-Aided Design* (CAD) software consisting of 20 pairs of 3D models, one representing the tumor and the other representing the organ for measuring the RCSA. The estimation of RCSA presented a median error of 2.01%(IQR 2.77%). The source code and the executable can be found at the following link github.com/ACarfi/contact-surface-areagui.

### 2.5 CSA measured with Hsieh’s method

In addition, we utilized the CT images to replicate the approximate formula proposed by Hsieh for evaluating the CSA values (HCSA) [13]. The formula is 2 ∗ *π* ∗ *r* ∗ *d* where *r* is the maximum radius of the tumor and *d* represents the maximum depth of the tumor into the uninvolved parenchyma considering the axes perpendicular to the tumor Figure 4. The CSAH was calculated in a double-blind manner by two radiologists, one with more than 20 years of experience in CT reporting, and the other with 3 years of experience. In our statistical analysis we focus on the measurement conducted by the radiology with more experience.

**Figure 4:**
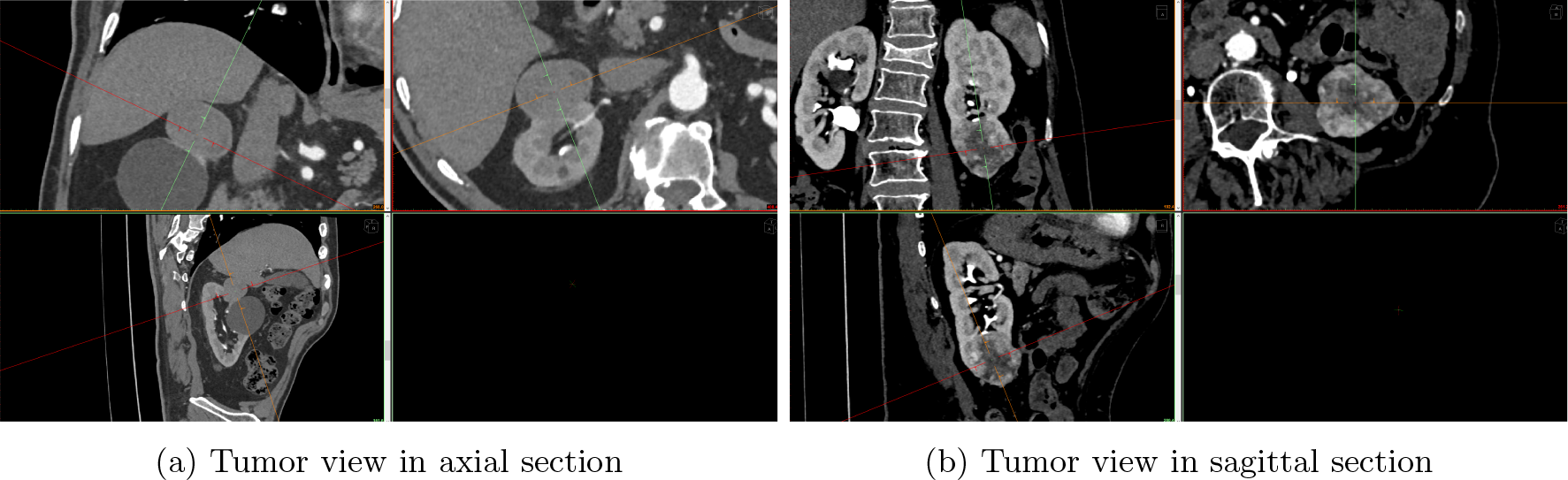
These images showcase the selection of sections for measuring the tumor’s maximum radius and maximum depth into the uninvolved parenchyma, which is contingent on the tumor’s shape, for area calculation using Hsieh’s method. In panel a, the axial section was opted for, while in panel b, the sagittal section was chosen.

### 2.6 Statistical Analysis

The statistical analysis was conducted using the software *IBM SPSS statistic* version *29*.*0*.*1*.*0*.. Continuous and ordinal variables are shown as median and interquartile range (IRQ), while categorical variables as percentage.

To determine the normality of the data, the *Kolmogorov-Smirnov* test was employed. Since the all data distribution was found to be non-normal, non-parametric test were used for analysis. Spearman’s correlation was utilized to investigate potential correlation between RCSA, HCSA, PADUA score, R.E.N.A.L. score and pre-operative, intra-operative and follow-up parameters. *Receiver Operating Charateristic* (ROC) curve were used to evaluate the performances of RCSA, PADUA score and R.E.N.A.L. score factors compared to intra-operative parameters such as WIT, OT, EBL. Furthermore, multivariable linear regression was used to determine if RCSA could predict intra-operative parameters. The criteria required for applying linear regression were tested prior to conducting the analysis.

A significance level of 0.05 was chosen, serving as the threshold for accepting or rejecting the hypotheses in the various tests.

## 3 Results

Of the 78 patients that underwent RAPN, 19 were female (24%) and 59 male (76%). As shown in the Table 1 the median age was 64 year (56-73 as IQR), BMI was 26 kg/m^2^ (23.88-29.41 as IQR). The median WIT was 17 min (11-22 IQR), OT was 180 min (130-210 IQR) and EBL was 100 mL (57.5-287.5 IQR). We obtained the following meadian and IRQ of the surface and tumor volume: RCSA was 14.37 cm^2^ (7.37-24.15), total area and total volume computed using the algorithm were 30.44 cm^2^ (16.13-66.23) and 14.24 cm^3^ (6.54-45.28) respectively, HCSA measured by the radiology with more experience was 15.02 cm^2^ (6.17-26.28). PADUA score had a median of 8,5 (7-10 IQR) while R.E.N.A.L. score 7 (5-8 IQR). Median values of pre-operative parameters were HB 13.85 g/dL (12.5-14.9 IQR), CREA 1.0 mg/dL (0.8-1.1 IQR), eGFR 82 mL/min/1.73m^2^ (64.25-93.25). The post-operative parameters had the following median values HB was 12.1 g/dL (11-13.6 IQR), CREA was 1.0 mg/dL (0.9-1.2 IQR) and eGFR was 75 mL/min/1.73m^2^ (57-91 IQR). Instead for the follow-up parameters we obtained HB 14 g/dl (12.63-15 IQR), CREA 1.1 mg/dL (0.9-1.2 IQR) and eGFR 73 mL/min/1.73m^2^ (61-87.5 IQR).

**Table 1:** Descriptive characteristics of study population.

|  | MEDIAN | IQR |
| --- | --- | --- |
| **Demographic data** |  |  |
| Age | 64 | 56-73 |
| BMI [kg/m<sup>2</sup>] | 26 | 23.88-29.41 |
| **Pre-operative clinical data** |  |  |
| HB [g/dL] | 13.85 | 12.5-14.9 |
| CREA [mg/dL] | 1 | 0.8-1.1 |
| eGFR [mL/min/1.73m<sup>2</sup>] | 82 | 64.25-93.25 |
| **Post-operative clinical data(3gg)** |  |  |
| HB [g/dL] | 12.1 | 11-13.6 |
| CREA [mg/dL] | 1 | 0.9-1.2 |
| eGFR [mL/min/1.73m<sup>2</sup>] | 75 | 57-91 |
| PCE | -0.1 | -0.13-0.05 |
| **Follow-up clinical data** |  |  |
| HB [g/dL] | 14 | 12.63-15 |
| CREA [mg/dL] | 1.1 | 0.9-1.2 |
| eGFR [mL/min/1.73m<sup>2</sup>] | 73 | 61-87.5 |
| **Operative Data** |  |  |
| WIT [min] | 17 | 11-22 |
| OT [min] | 180 | 130-210 |
| EBL [mL] | 100 | 57.5-287.5 |
| PADUA | 8.5 | 7-10 |
| R.E.N.A.L. | 7 | 5-8 |
| **Dimensional tumor characteristics** |  |  |
| Total Volume [cm<sup>3</sup>] | 14.24 | 6.54-45.28 |
| Total Area [cm<sup>2</sup>] | 30.44 | 16.13-66.23 |
| RCSA [cm<sup>2</sup>] | 14.37 | 7.37-24.5 |
| HCSA [cm<sup>2</sup>] | 15.02 | 6.17-26.28 |

IRQ = interquartile range BMI = Body Mass Index HB = Haemoglobin CREA = Creatinine eGFR = estimate glomerular filtration rate PCE = percentage change in eGFR WIT = Warm Ischemia Time OT = Operative Time EBL = Estimate Blood Loss RCSA = Real Contact Surface Area HCSA = Hsieh's Contact Surface Area

As can be seen in Table 2 the Spearman’s test showed a correlation between RCSA and intra-operative parameters. There was no correlation between RCSA and pre-operative parameters. Instead, there was a direct correlation with the CREA post-operative (*ρ* = 0.27 & p = 0.01) and an inverse correlation with eGFR post-operative (*ρ* = -0.32 & p = 0.004). Furthermore, we found a relation also with the follow-up parameters CREA (*ρ* = 0.38 & p = 0.004) and eGFR (*ρ* = -0,34 & p = 0.01). The correlation between RCSA and HB of follow-up turned out to be not significant (p-value*>*0.05).

**Table 2:** Speraman’s Correlation.

|  | RCSA |  | HCSA |  | PADUA |  | R.E.N.A.L. |  |
| --- | --- | --- | --- | --- | --- | --- | --- | --- |
| | $\rho$ | P | $\rho$ | P | $\rho$ | P | $\rho$ | P |
| RCSA | 1.000 | - | 0.9 | <0.001 | 0.53 | <0.001 | 0.52 | <0.001 |
| HCSA | 0.9 | <0.001 | 1.000 | - | 0.52 | <0.001 | 0.53 | <0.001 |
| PADUA | 0.53 | <0.001 | 0.52 | <0.001 | 1.000 | - | 0.64 | <0.001 |
| R.E.N.A.L. | 0.52 | <0.001 | 0.53 | <0.001 | 0.64 | <0.001 | 1.000 | - |
| WIT | 0.64 | <0.001 | 0.64 | <0.001 | 0.27 | 0.01 | 0.47 | <0.001 |
| OT | 0.47 | <0.001 | 0.44 | <0.001 | 0.27 | 0.04 | 0.24 | <0.03 |
| EBL | 0.57 | <0.001 | 0.61 | <0.001 | 0.28 | <0.01 | 0.29 | <0.01 |
| **Pre-operative data** |  |  |  |  |  |  |  |  |
| eGFR | -0.09 | 0.4 | -0.03 | 0.7 | 0.13 | 0.2 | -0.11 | 0.3 |
| CREA | 0.07 | 0.4 | -0.41 | 0.7 | 0.06 | 0.5 | 0.12 | 0.3 |
| HB | -0.43 | 0.7 | -0.01 | 0.8 | -0.12 | 0.1 | -0.21 | 0.06 |
| **Post-operative data** |  |  |  |  |  |  |  |  |
| eGFR | -0.32 | 0.004 | -0.29 | 0.01 | -0.26 | 0.02 | -0.2 | 0.17 |
| CREA | 0.27 | 0.01 | 0.28 | 0.01 | 0.2 | 0.06 | 0.19 | 0.09 |
| HB | -0.13 | 0.2 | -0.13 | 0.26 | -0.12 | 0.2 | -0.16 | 0.1 |
| PCE | -0.2 | 0.07 | -0.19 | 0.09 | -0.07 | 0.5 | -0.4 | 0.6 |
| **Follow-up data** |  |  |  |  |  |  |  |  |
| eGFR | -0.34 | 0.01 | -0.2 | 0.04 | -0.16 | 0.23 | -0.26 | 0.05 |
| CREA | 0.38 | 0.004 | 0.32 | 0.01 | 0.12 | 0.3 | 0.25 | 0.05 |
| HB | -0.13 | 0.3 | -0.13 | 0.3 | -0.04 | 0.7 | -0.29 | 0.03 |

RCSA = Real Contact Surface Area HCSA = Hsieh's Contact Surface Area WIT = Warm Ischemia Time OT = Operative Time EBL = Estimate Blood Loss eGFR = estimate glomerular filtration rate CREA = Creatinine HB = Haemoglobin PCE = percentage change in eGFR

Also, HCSA, as shown in previous works, was correlated with intra-operative parameters and follow-up parameters. PADUA score was correlated with intra-operative parameter, post-operative eGFR but had no correlation with followup parameters.

R.E.N.A.L. score exhibits a strong correlation with WIT (*ρ* = 0.47 & p *<* 0.001), OT (*ρ* = 0.25 & p = 0.03) and EBL (*ρ* = 0.29 & p = 0.01). R.E.N.A.L. was also correlated with follow up HB (*ρ* = -0.29 & p = 0.03), but there was no correlation with follow-up eGFR (p*>*0.05). We further examined the correlation between RCSA and HCSA, PADUA score, and R.E.N.A.L. score. The PCE was found to be unrelated to both scores and the two CSA.

We conducted an analysis, based on ROC curve to evaluate the performance of the RCSA, PADUA score and R.E.N.A.L. score in comparison to intra-operative parameters. In all three cases, our parameter yielded the better results. For each of the intra-operative parameters (OT,WIT,EBL), we selected a value against which we evaluated the three factors. Specifically, we opted for a threshold of 180 minutes for OT, which represents its median value. As for WIT, we chose 20 minutes as the maximum allowable time, considering that exceeding this threshold could adversely impact renal function [14, 15]. In the case of EBL, we selected a limit of 200 mL because it was established as the cut-off value for the transperitoneal RAPN [16], the surgical method employed in our cases.

The *Area Under the Curve* (AUC) is a single scalar values that measure the performance of RCSA, PADUA and R.E.N.A.L. score in ROC analysis. It was shown that RCSA was a better predictor for WIT (AUC = 0.84 vs 0.66 vs 0.71), OT (AUC = 0.75 vs 0.62 vs 0.63) and EBL (AUC = 0.72 vs 0.62 vs 0.64) than PADUA and R.E.N.A.L. score as can be seen in the Figure 5.

**Figure 5:**
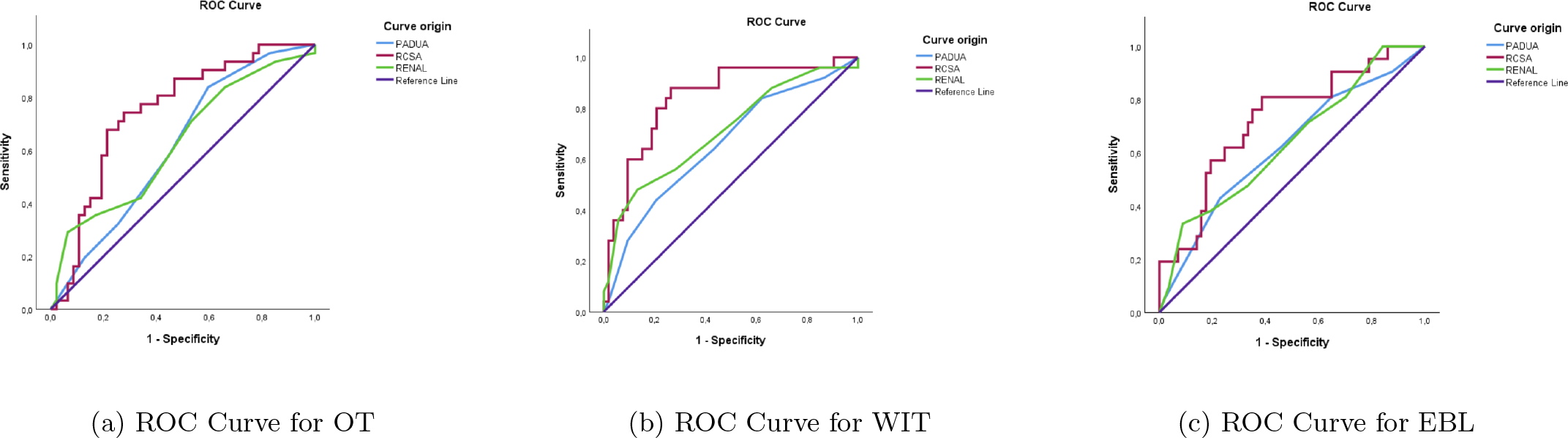
Reported here are the ROC Curves of each of the three intra-operative parameters.

Besides AUC can be derived from ROC curve also a cut-off values for the three parameters, computed using *Youden’s Index*. The RCSA cut-off values for OT and WIT is 15 cm^2^ while for EBL is 14 cm^2^.

We also performed multivariable linear regression to demonstrate that RCSA is a significant independent predictor of intra-operative parameters (Table 3). The independent variables considered in the regression were RCSA, PADUA score and R.E.N.A.L. score. Consistent with the findings from the ROC curves, RCSA exhibited good predictive capability.

**Table 3:** Multivariable Linear Regression analyses testing the association between RCSA, PADUA and R.E.N.A.L. and WIT, OT and EBL.

|  | WIT |  |  | OT |  |  | EBL |  |  |
| --- | --- | --- | --- | --- | --- | --- | --- | --- | --- |
|  | B | 95% CI | p | B | 95% CI | p | B | 95% CI | p |
| RCSA | 0.39 | (0.1 - 0.6) | 0.003 | 0.35 | (0.07 - 0.6) | 0.01 | 0.48 | (0.2 - 0.7) | <0.001 |
| PADUA | -0.22 | (-0.5 - 0.05) | 0.11 | -0.02 | (-0.3 - 0.2) | 0.8 | -0.1 | (-0.4 - 0.18) | 0.46 |
| R.E.N.A.L. | 0.36 | (0.09 - 0.6) | 0.01 | 0.07 | (-0.22 - 0.36) | 0.6 | -0.01 | (-0.3 - 0.27) | 0.9 |

RCSA = Real Contact Surface Area WIT = Warm Ischemia Time OT = Operative Time EBL = Estimate Blood Loss

These regression results support the notion that RCSA is a strong and significant predictor of WIT, OT, and EBL in our study.

## 4 Discussion

In our study, similar to previous research [11, 17], we addressed the correlation between peri-operative parameters (i.e., WIT, OT, and EBL) and RCSA, PADUA score and R.E.N.A.L. score. We discovered a significant correlation between these factors and with intra-operative parameters. Furthermore, by employing ROC curve analysis, we determined that RCSA outperformed both PADUA score and R.E.N.A.L. score as a predictive factor for each intra-operative parameter. We also obtained through Youden’s index cut-off values of RCSA respect all the three intra-operative parameters: for WIT and OT the cut-off values was 15 cm^2^ while for EBL was 14 cm^2^.

Moreover, our findings indicate that RCSA is a reliable independent predictor for WIT, OT, and EBL. Also R.E.N.A.L. score was found to be an independent predictor for WIT. These results highlight the superior predictive ability of RCSA over other scoring systems, emphasizing its potential value in guiding surgical decision-making and improving patient outcomes.

The concept of CSA was initially developed by Leslie et al. [10]. In their work, CSA was proposed as a predictor of eGFR function. However the method they used to estimate CSA was incorrect. They computed the total area of the tumor by approximating it as a sphere and determining the percentage of intraparenchymal components. This method was discredited by Hsieh et al. [13] in 2016 with the subsequential proposal of a new mathematical formula 2 ∗ *π* ∗ *r* ∗ *d* for evaluating CSA. However, even in Hsieh’s work, the tumor was still approximated as a sphere, which had its limitations as mentioned by authors themselves in the study.

The assumption that tumors are spherical holds little relevance in our case study, primarily consisting of RCC, tumors characterized by a highly variable morphology, so the Hsieh’s formula has a restrictive applicability (Figure 6).

**Figure 6:**
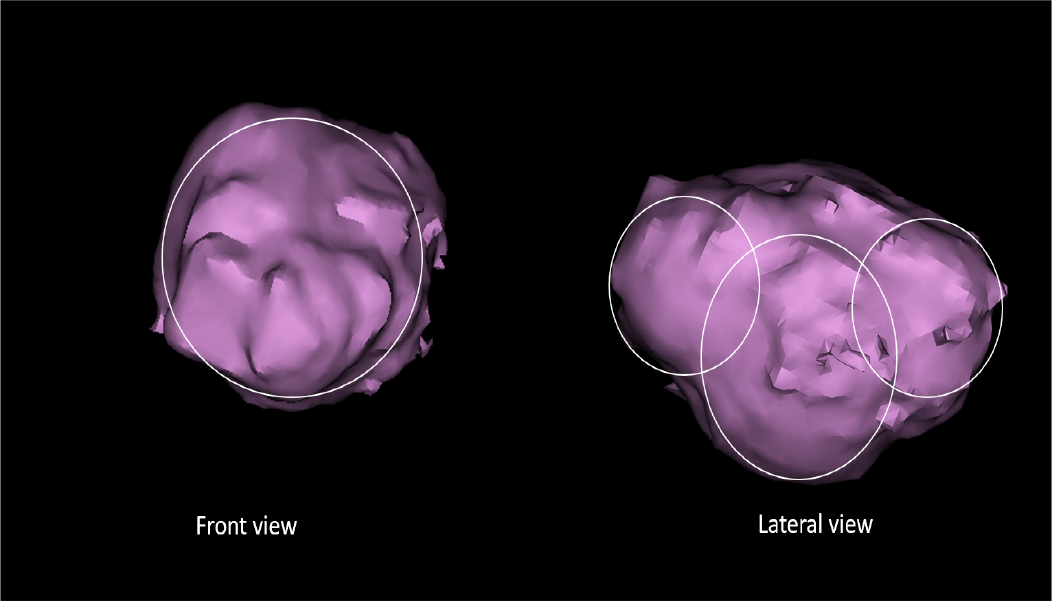
This image illustrates the potential confusion in interpretation. In the front view, the tumor appears to be approximated as a sphere, while in the lateral view, it seems to be comprised of three distinct spheres.

In addition, the method proposed by Hsieh has a significant drawback which consists of its operator-dependency. The visual selection of parameters required to calculate the contact area within medical images is closely tied to the operator conducting the measurement. To address this limitation, we introduce an alternative approach based on 3D reconstructions and an algorithm designed to compute the area based on the actual geometry of the reconstructed 3D object. This method demonstrates high accuracy, as evidenced by the minimal error produced during objective tests used to validate the algorithm’s functionality.

Nevertheless, the current manual segmentation of the 3D models for the kidney and tumor introduces an element of potential inaccuracy, as even an experienced operator cannot ensure consistent reproducibility. This concern, however, can be alleviated by adhering to precise parametric standards during segmentation, which are connected to the Hounsfield units of the radiological image. Furthermore, technological advancements are anticipated to enable automated machine segmentation, a capability already present for certain organ types (e.g., as demonstrated by Materialise Mimics in the case of the heart). Looking ahead, we anticipate an automated reconstruction of organs and tumors with maximum reproducibility. This, in combination with our algorithm, will facilitate a highly accurate calculation of the contact area, enhancing the realism of the results.

Leslie et al. [10] suggested a cut-off value of 20 cm^2^ for CSA to estimate differences in peri-operative parameters. However, the specific methods they used to determine this cut-off value were not provided in their work. Ficarra et al.[18], in their study, adopted the same cut-off value of 20 cm^2^ proposed by Leslie et al. [10] to investigate the differences in peri-operative parameters using this threshold.

In our study, we employed ROC curve analysis to evaluate the effectiveness of our method. By examining the sensitivity and specificity values, we determined the Youden’s index, which allowed us to establish cut-off values for RCSA in relation to the three major intra-operative parameters. For OT and WIT, we obtained a cut-off value of 15 cm^2^, while for EBL, the cut-off value was 14 cm^2^. These values are lower than the cut-off value proposed by Leslie et al., but we believe they are more realistic and provide better discrimination in predicting the peri-operative parameters as calculated with a system adhering to 3D reconstruction.

Other studies conducted after Hsieh’s work have also explored CSA estimation using 3D reconstruction techniques. Bianchi et al. [19] utilized Autodesk MeshMixer software for reconstruction purposes, but they did not propose a specific method for CSA calculation. They found that the contact surface area was a predictor of WIT.

Takagi et al. [20] proposed the segmentation of the kidney and tumor using Materialise Mimics software, and CSA was calculated by “hand-drawing” on the 3D reconstructed kidney and automatically measured by the medical software. It is, however, an arbitrary method and not reproducible without an error to calculate. They found the CSA to be a predictor of the decrease rate of eGFR and to be associate with the decrease of operated parenchyma.

Umemoto et al. [21] proposed organ segmentation using the software Synapse, and after reconstruction, they simulated the removal of the tumor and measured the area of contact as the parenchymal area removed. However, their method lacks a proper estimation technique for accurate CSA values, and considering CSA as the area post-surgery is not a correct measure. Furthermore, their study was retrospective.

In contrast, our work utilized 3D reconstruction of the kidney and tumor using Materialise software. The segmentation of organs in our study is based on DICOM images obtained from our innovative protocol. This protocol was specifically designed to capture all phases (arterial, venous, late) in a single acquisition. By utilizing this protocol, we ensure comprehensive imaging coverage and enable more accurate segmentation of the organs of interest. We developed a dedicated algorithm that precisely computes the values of the RCSA, taking into account the geometric features of the reconstructed kidney and tumor. This approach ensures a more accurate and reliable measurement of CSA.

The algorithm we developed exhibits high reproducibility and is accessible to anyone with STL files of organs and tumors. The link provided in the materials and methods section allows users to access and utilize the algorithm for their own research or clinical purposes, thereby maintaining anatomical measurements’ correspondence and enabling evaluation of pre-operative strategies and predictability.

However, our study is not devoid of limitations. First, the study population is limited in size. Therefore, larger series are warranted in order to enhance results’ robustness and reliability. Second, a single highly-experienced robotic surgeon performed all the surgeries. In consequence, our data might not be generalized to other centers with lower case-load and expertise. Last but not least, our data lack of validation. Therefore, further external validations from additional data sources are recommended to corroborate our findings.

## 5 Conclusions

3D reconstruction of renal, tumor and vascular anatomy before RAPN could be a useful tool to predict peri-operative outcomes during and after surgery. The ability to overlay the reconstruction on top of the robot viewer using during operations also aids the surgeon. Based on this reconstruction we measured the *Real Contact surface Area* which has been demonstrated to be a good predictor of peri-operative outcomes.

Further studies are needed in order to confirm and validate our results.

## Data Availability

All data generated for this analysis were from anonymized database. The code for the analyses will be made available upon request.

